# Multi-omic network analysis identifies dysregulated neurobiological pathways in opioid addiction

**DOI:** 10.1101/2024.01.04.24300831

**Authors:** Kyle A. Sullivan, David Kainer, Matthew Lane, Mikaela Cashman, J. Izaak Miller, Michael R. Garvin, Alice Townsend, Bryan C. Quach, Caryn Willis, Peter Kruse, Nathan C. Gaddis, Ravi Mathur, Olivia Corradin, Brion S. Maher, Peter C. Scacheri, Sandra Sanchez-Roige, Abraham A. Palmer, Vanessa Troiani, Elissa J. Chesler, Rachel L. Kember, Henry R. Kranzler, Amy C. Justice, Ke Xu, Bradley E. Aouizerat, VA Million Veteran Program, Dana B. Hancock, Eric O. Johnson, Daniel A. Jacobson

**Author notes:** This manuscript has been authored by UT-Battelle, LLC under Contract No. DE-AC05-00OR22725 with the U.S. Department of Energy. The United States Government retains and the publisher, by accepting the article for publication, acknowledges that the United States Government retains a non-exclusive, paid-up, irrevocable, world-wide license to publish or reproduce the published form of this manuscript, or allow others to do so, for United States Government purposes. The Department of Energy will provide public access to these results of federally sponsored research in accordance with the DOE Public Access Plan (http://energy.gov/downloads/doe-public-access-plan).

## Abstract

Opioid addiction constitutes a public health crisis in the United States and opioids cause the most drug overdose deaths in Americans. Yet, opioid addiction treatments have limited efficacy. To help address this problem, we used network-based machine learning techniques to integrate results from genome-wide association studies (GWAS) of opioid use disorder and problematic prescription opioid misuse with transcriptomic, proteomic, and epigenetic data from the dorsolateral prefrontal cortex (dlPFC) in opioid overdose victims. We identified 211 highly interrelated genes identified by GWAS or dysregulation in the dlPFC of individuals with opioid overdose victims that implicated the Akt, BDNF, and ERK pathways, identifying 414 drugs targeting 48 of these opioid addiction-associated genes. This included drugs used to treat other substance use disorders and antidepressant drugs. Our synthesis of multi-omics using a systems biology approach revealed key gene targets that could contribute to drug repurposing, genetics-informed addiction treatment, and future discovery.

## Introduction

Worldwide, over 60 million people misuse opioid drugs resulting in an estimated 12.9 million healthy years of life lost (disability-adjusted life years)^1^. These drugs include both opioids prescribed for pain relief (*e.g.*, hydrocodone, oxycodone) and illicit opioids (*e.g.*, heroin, fentanyl). In the United States, over 9 million people in the housed population (which excludes homeless and institutionalized individuals) aged 12 and older were estimated to misuse opioids in 2020^2^, and over 80,000 people died of an opioid-related overdose in 2021 – continuing a decades long epidemic of opioid misuse and overdose deaths^3,4^. Yet only 1.27 million people in the United States receive medication-assisted treatment with one of the three medications approved to treat opioid use disorder (OUD)^5^: methadone (opioid agonist), buprenorphine (opioid agonist/antagonist), or naltrexone (opioid antagonist). Substance use disorders are widely accepted to involve genetic and experiential influences on brain circuits related to motivated behavior. Thus, there is a great need for a better understanding of the neurobiology of opioid addiction and the identification of novel targets for drug development.

Several recent genome-wide association studies (GWAS)^6–10^ have identified genetic variants and genes associated with increased risk of opioid addiction phenotypes. Other studies have identified gene dysregulation in postmortem human brains associated with opioid overdose deaths^11–15^. Up to 18 genome-wide significant loci have been reported across the most recent GWASs for OUD or prescription opioid misuse, with replicated associations being observed for *OPRM1* ^7–10^, *FURIN* ^7–9^, the *SCAI/PPP6C/RABEK* cluster ^7–9^, and *PTPRF* ^6,9^. In parallel, four RNA-seq studies identified hundreds of potentially differentially expressed genes in human postmortem dorsolateral prefrontal cortex (dlPFC), which were enriched for a variety of biological functions (*e.g.,* extracellular matrix, angiogenic cytokines, and MAPK signaling). These studies did not include replication, but we found that 11 genes replicate across these cohorts for the dlPFC (Bonferroni corrected *p* < 0.05), and a meta-analysis^16^ of these cohorts identified up to 335 dysregulated genes in the dlPFC (Benjamini-Hochberg FDR p-value < 0.05).

Although these recent findings provide important clues to the biology underlying opioid addiction, these genes do not function in isolation. Additional insights may be gained through a systems biology approach that identifies affected functional networks from disease-associated genes^17^. Here, we used results from existing studies to identify 404 opioid addiction-related genes from which we removed potential false positives with the recently developed Gene set Refinement through Interacting Networks (GRIN^17^) software. GRIN enabled us to identify a tightly integrated set of 211 genes that mapped to multiple neurobiological pathways. Fifty of the 211 genes implicated in opioid addiction signaling or other substance use disorders were tightly interconnected using cross validation and a concise shortest-paths network between pairs of genes. We created a conceptual model of the network mapped to BDNF and MAPK signaling pathways and synaptic signaling processes, among others, showing widespread downregulation of these genes in opioid addiction. Moreover, multiple genes appear to be promising targets for novel drug repurposing for treating OUD based on their role as gene targets for drugs used to treat other substance use disorders. Our results demonstrate the utility of integrating multiple omics in systems biology approaches that leverage machine learning techniques, discovering novel biological relationships that underlie opioid addiction.

## Results

### Meta-analysis of opioid addiction omics data sets combined with network biology identifies 211 highly interrelated genes

We first identified opioid addiction-associated genes from multiple omics data sets using the postmortem dlPFC and GWAS SNP-nearest gene assignment (**Figure 1**). Using consistent significance thresholds within each omics data set across studies (Online Methods), we identified 404 unique opioid addiction genes from the following omics data types: 256 genes associated with H3K27ac ChIP-seq peaks^11^, 13 DNA methylation-associated genes^15^, 33 GWAS-associated genes^6–10^, 3 protein-coding genes associated with differentially abundant proteins from LC/MS proteomics^12^, and 104 differentially expressed genes from RNA-seq^11–14^ (**Supplementary Tables 1-7**).

**Figure 1.**
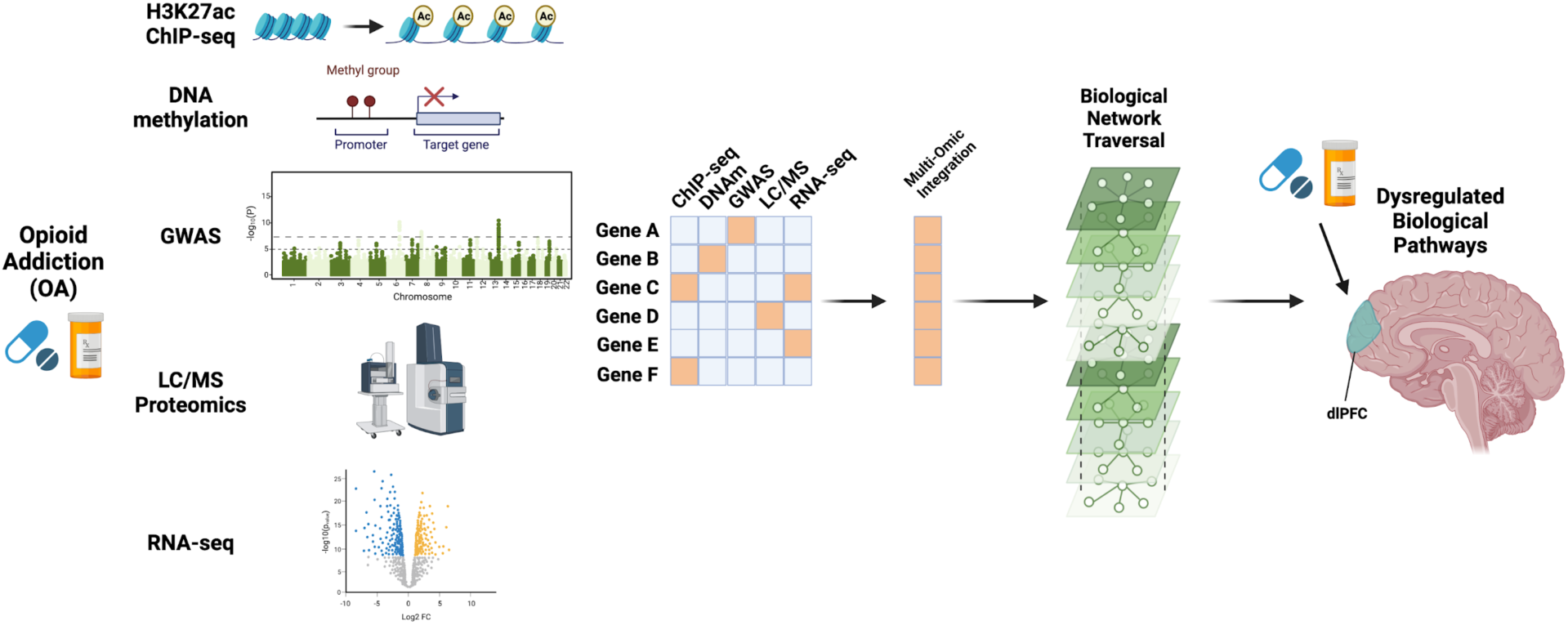
Multi-omic integration of opioid addiction phenotypes. Beginning with multiple data sets collected from opioid addiction (OA) cases and controls, OA genes were assembled from H3K27ac chromatin immunoprecipitation (ChIP-seq; peaks assigned to nearest gene), DNA methylation (CpG methylation site to nearest gene), genome-wide association studies (GWAS; SNP-to-gene assignment), differentially abundant proteins by LC/MS proteomics (protein-coding genes), and differential gene expression by RNA-seq. After integrating the overlapping and distinct genes identified by each omics data type, a biological multiplex network consisting of networks from multiple lines of biological evidence were constructed using data sources separate from any of the opioid addiction omics data sets. Network traversal algorithms were used to identify mechanistic connections among the multi-omic genes and identify dysregulated pathways in the dorsolateral prefrontal cortex (dlPFC). Figure made with Biorender.com.

Of the 404 opioid addiction-related genes, we sought to identify those that are highly interconnected in biological networks using multiple lines of experimental evidence from data independent of the opioid omics data sets. Using GRIN^17^, we removed potential false positive genes based on network connectivity within a multiplex network consisting of 10 layers of biological evidence. GRIN uses the algorithm Random Walk with Restart (RWR) to identify tightly interconnected genes and removes those that do not depart from the gene ranks of a null distribution. GRIN identified 211 highly interrelated opioid addiction genes across omics data types (**Figure 2A, Supplementary Tables 8-9**).

**Figure 2.**
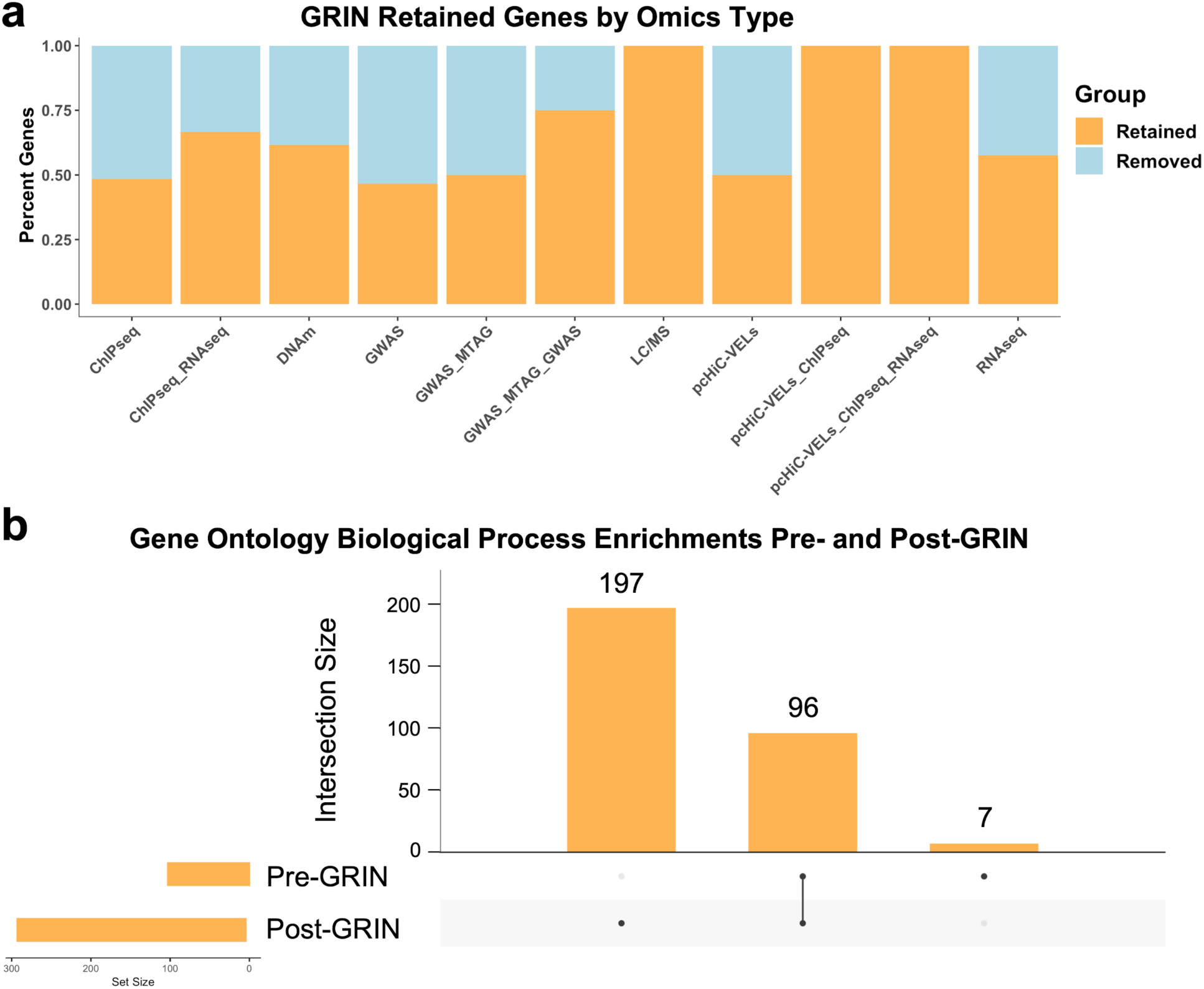
Opioid addiction genes are retained by GRIN from multiple omics types and result in more Gene Ontology enrichments compared to the unfiltered gene set. **a.** Percentages of genes retained (orange) or removed (blue) by GRIN by each omics type, including genes shared across omics types (ChIP-seq_RNAseq, GWAS_MTAG_GWAS, pcHiC-VELs_ChIPseq, and pcHiC-VELs_ChIPseq_RNAseq). **b.** From the 404 original opioid addiction genes (Pre-GRIN), 159 GO Biological Process terms were significantly enriched. The 211 genes retained by GRIN (Post-GRIN) were enriched for 96 of the same GO Biological Processes but were uniquely enriched for 197 additional terms. Only 7 terms were significantly enriched in the set of 404 genes prior to GRIN that were not significantly enriched in the post-GRIN set of 211 genes.

Before GRIN filtering, the 404 unfiltered genes were significantly enriched for 103 Gene Ontology (GO^18^) Biological Process terms. The 211 genes that remained after GRIN filtering were enriched for 293 terms, including 96 of the same enriched biological processes as were in the unfiltered gene set prior to the application of GRIN (**Figure 2B, Supplementary Tables 10-11**). In contrast, the 193 genes removed by GRIN were not significantly enriched for any GO Biological Process terms, clearly differentiating them from the 211 retained genes. Thus, the subset of 211 genes retained by GRIN were highly biologically interrelated based on interconnectivity within the biological multiplex network and better mapping to known biological processes.

### A subset of multi-omic genes previously implicated in opioid signaling and substance use disorders are highly interconnected in biological networks

Of the 211 genes retained by GRIN, we focused on a subset of 50 unique, high-confidence genes (**Supplementary Table 12**) that were either implicated across 2 or more studies or previously implicated in opioid signaling or in other substance use disorders (Online Methods). Gene set enrichment analyses indicated that, among other functions, these high confidence genes were enriched for the BDNF and MAPK signaling pathways and synaptic signaling processes (**Supplementary Table 13**).

Using shortest paths network traversal to identify the fewest connections between each pair of the 50 high-confidence opioid addiction genes, we found that 43 were directly connected to at least one other gene in the set (**Figure 3A, Supplementary Table 14**). Moreover, the other 7 high confidence genes were only two connections away from at least one of the other 50 high confidence genes (**Figure 3A, Supplementary Table 14**). Only 127 additional network-connecting genes (not high-confidence opioid addiction genes) were necessary to link pairs of genes that could be connected as direct neighbors or by a shared neighboring gene. Furthermore, 3 of the 127 network-connecting genes (*SERPINB1*, *SORCS1*, and *SORL1*) were members of the original 211 GRIN-retained gene set (**Figure 3A, Supplementary Table 15**).

**Figure 3.**
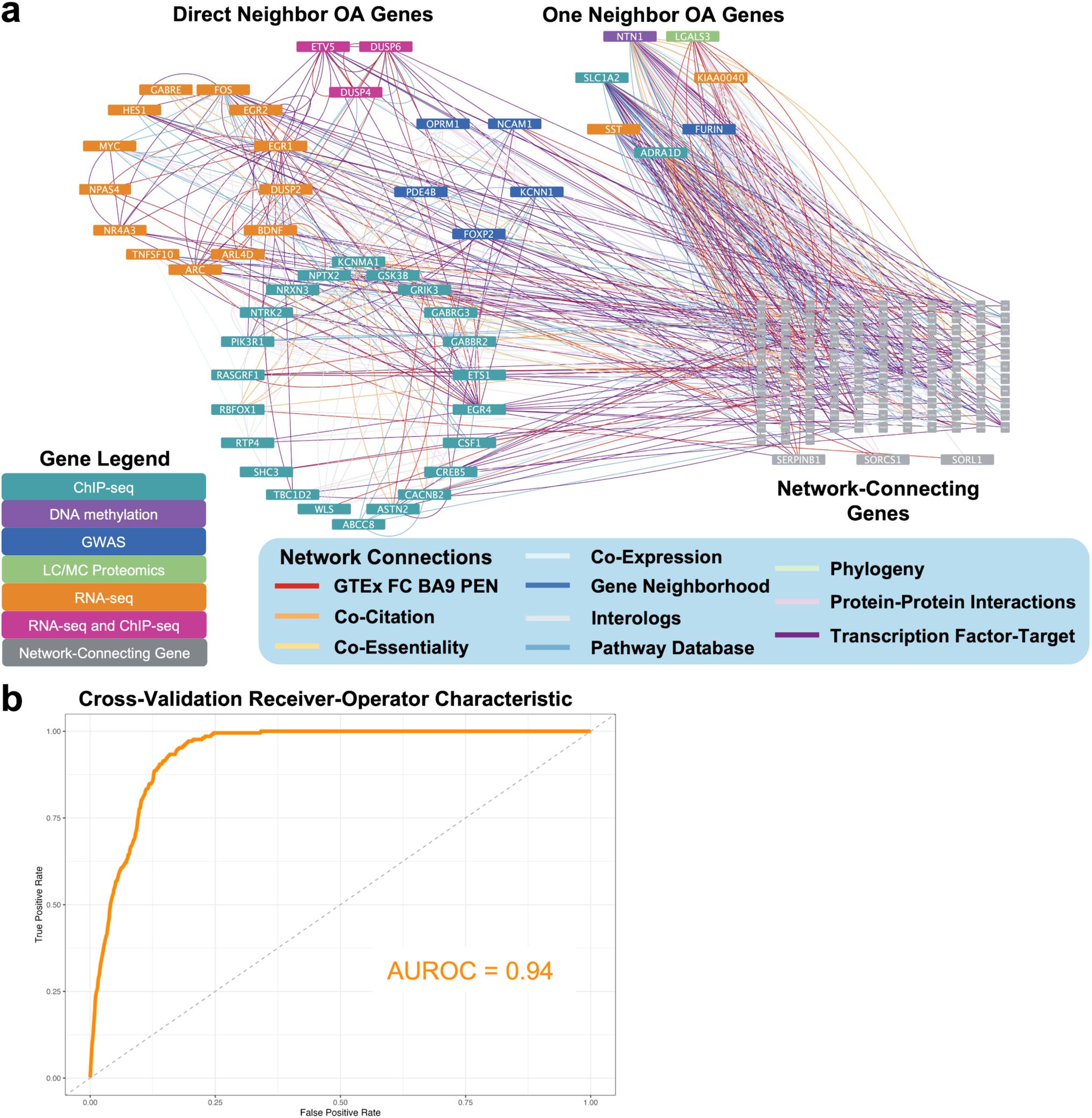
Multi-omic opioid addiction genes are tightly interconnected as demonstrated by network biology. **a.** Network visualization of the shortest pathways between all pairs of 50 opioid addiction (OA)-associated genes from GWAS and dlPFC omics. 43 genes were directly connected to at least one other gene by the networks (Direct Neighbor OA genes), and only 127 additional genes (Network-Connecting Genes) were necessary to connect the other 7 genes (One Neighbor OA Genes) from 10 network layers. Notably, three Network-Connecting genes (*SERPINB1, SORCS1,* and *SORL1*) were members of the larger 211 GRIN-retained opioid addiction gene set. Gene Legend: indicates gene color from which omics data type or if it is a Network-Connecting Gene; Network Connections legend indicates network layer used to connect gene pairs. **b.** Using Random Walk with Restart to explore the biological networks starting from 50 OA-associated genes, 5-fold cross validation exhibits high recall based upon a mean area under receiver-operator characteristic curve (AUROC) value of 0.94.

Next, we explored the interconnectivity of the 50 high-confidence opioid addiction genes using RWR. We employed a cross-validation approach in which a subset of the opioid addiction genes were used as starting points to explore the biological networks with RWR, and the remaining genes were ranked against the rest of the genes in the biological network based on how often the gene was visited by RWR. Using this cross-validation approach, we observed a high area under receiver-operator characteristic curve (AUROC) value (AUROC = 0.94; **Figure 3B**). The high AUROC value indicated that, during cross validation, genes that were used to explore the networks frequently visited the genes that were left out of the gene set. Combined with our small shortest paths network, these results confirmed that these 50 genes were highly interconnected in our biological multiplex network.

### Conceptual model of biological pathways underlying opioid addiction multi-omic genes

After integrating the 211 multi-omic opioid addiction genes using our network approaches, we identified multiple pathways unifying these genes. From this investigation, we developed a conceptual model consisting of 45 opioid addiction genes and 26 other genes, proteins, or molecules in associated pathways (**Figure 4**). Multiple genes in the BDNF pathway were implicated, including *BDNF*, whose expression was downregulated and its receptor *NTRK2* (TrkB), which exhibited decreased H3K27ac ChIP-seq peaks in opioid samples (**Figure 4**). A number of genes were also implicated downstream of the BDNF pathway, including *RASGRF1*, *PIK3R1* and *GSK3B* in the PI3K/Akt pathway. The ERK MAPK pathway was also strongly implicated, as the μ-opioid receptor (*OPRM1*), GABA_B_ receptor (*GABBR2*), and *PDE4B* were implicated upstream of ERK along with multiple ERK phosphatases (*DUSP2*, *DUSP4*, *DUSP6*, *DUSP10*, *PPP6C*). Downstream of the ERK and Akt pathways, CREB (*CREB5*) and a number of its target genes were also implicated in opioid addiction, including several immediate early genes (*ARC*, *EGR1*, *EGR2*, *EGR4*, *FOS*, *MYC*, *NPAS4*) and genes involved in synaptic plasticity (*BDNF*, *NTN1*)^19^. Additionally, the transcription factor *RORA* was predicted to activate the transcription of the multi-omic opioid addiction genes: astrotactin-2 (*ASTN2*) and galectin-3 (*LGALS3*)^20^.

**Figure 4.**
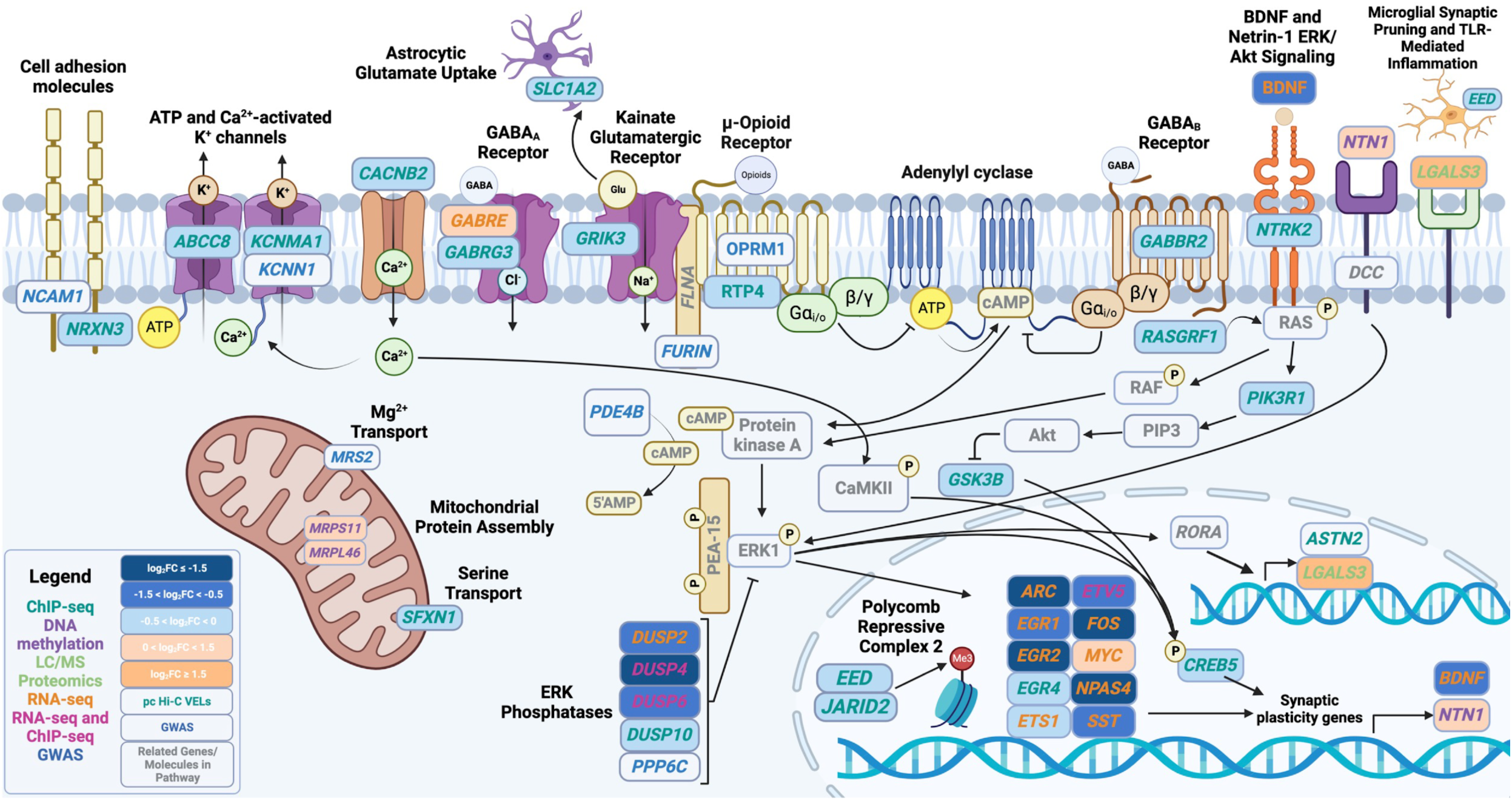
Conceptual model of opioid addiction pathways implicated by multi-omic integration. Conceptual model of 45 opioid addiction genes identified via multiple omics data types and 26 additional genes, proteins, or molecules in associated pathways. The mu-opioid receptor (*OPRM1*) and *GABBR2* receptor inhibit downstream adenylyl cyclase/protein kinase A (PKA) signaling. PKA can phosphorylate ERK1 which is bound by the scaffolding protein PEA-15, and ERK can also be activated by upstream netrin (*NTN1*) and BDNF signaling molecules that were implicated in opioid addiction (*BDNF*, *NTRK2*, *RASGRF1*). *DUSP2, DUSP4*, *DUSP6*, *DUSP10*, and *PPP6C* all function as ERK phosphatases and *PDE4B* can reduce PKA activation, while Akt signaling (implicated by *PIK3RA* and *GSK3B*) and ERK signaling can activate *NPAS4* and *CREB5* to activate transcription of *NTN1* (identified by DNA hypermethylation) and synaptic plasticity and immediate early genes (*ARC*, *BDNF, EGR1, EGR2, EGR4, ETS1, ETV5, FOS, MYC, NPAS4, SST*). ERK can also activate the transcription factor *RORA* to promote transcription of *ASTN2* and galectin-3 (*LGALS3*), which is important in microglial inflammatory processes. *OPRM1* (chaperoned to the cell membrane by *RTP4*) and *FURIN* share a common scaffolding protein (filamin A; *FLNA*) with the glutamatergic kainate receptor subunit *GRIK3*, and *SLC1A2* is an important glutamate transporter in astrocytes. Additional potassium and calcium channel subunits (*ABCC8*, *KCNMA1*, *KCNN1*) were implicated along with multiple ionotropic GABA_A_ receptor subunits (*GABRE*, *GABRG3*) and cell adhesion molecules (*NCAM1* and *NRXN3*). Color of gene text indicates which opioid addiction omics data set the gene originated from, and shading of the gene indicates the logFC of expression or histone acetylation state (not applicable for pc Hi-C VELs or GWAS genes). *NTN1* was hypermethylated by a mean difference of 0.29 rather than a logFC difference in methylation state. Gray text genes indicate genes or molecules involved in pathways but were not implicated by an omics study.

In addition to the BDNF, ERK, and Akt signaling pathways, multiple ion channel subunits and cell adhesion molecules that may influence neurotransmission were identified by multi-omic integration. Filamin A (*FLNA*) binds OPRM1 ^21^, FURIN ^22^, and the GRIK3 subunit of the kainate glutamatergic receptor^23^ (**Figure 4**). Moreover, the astrocytic glutamate transporter EAAT2 gene (*SLC1A2*) was implicated by hypoacetylated ChIP-seq. Multiple ionotropic GABA receptor subunit genes (*GABRE*, *GABRG3*), a voltage-gated calcium channel subunit gene (*CACNB2*), and multiple ATP or Ca^2+^-activated K^+^ channel subunit genes (*ABCC8*, *KCNMA1*, *KCNN1*) were identified by ChIP-seq, GWAS, or RNA-seq. Genes encoding cell adhesion molecules (*NCAM1* ^24^, *NRXN3* ^25^) that can modulate synaptic connectivity were also implicated in our multi-omic opioid addiction gene set, and microglial inflammation was implicated by the galectin-3 gene (*LGALS3* ^26^). Notably, most genes exhibited decreased gene expression, hypoacetylated H3K27ac peaks, or increased DNA methylation, except for increased *GABRE* and *MYC* gene expression and increased protein abundance of *LGALS3*.

### Putative pharmacological target genes in opioid addiction

After identifying biological pathways and highly interrelated genes by integrating the multi-omic opioid addiction data sets, we identified genes whose protein products are candidates for pharmacological manipulation and potential drug repurposing. We constructed a network of 48 druggable opioid addiction genes and 414 approved or experimental drugs known to target the products of these genes (**Figure 5, Supplementary Table 16**). Currently approved treatments for opioid use disorder (OUD) or overdose that target the mu-opioid receptor (encoded by *OPRM1*) are included in the network (buprenorphine, methadone, nalmefene, naloxone, and naltrexone). Naltrexone is also approved for treating alcohol use disorder (AUD)^27^, and other approved or investigational AUD treatments^27^ were also present in the network, such as acamprosate^27^ (*GABRE* and *GABRG3*), baclofen^27^ (*GABBR2*), ibudilast^28^ (*PDE4B*), and topiramate^27,29^ (*CACNB2*, *GRIK3*, and *SCN8A*; **Figure 5**, **Supplementary Table 16**).

**Figure 5.**
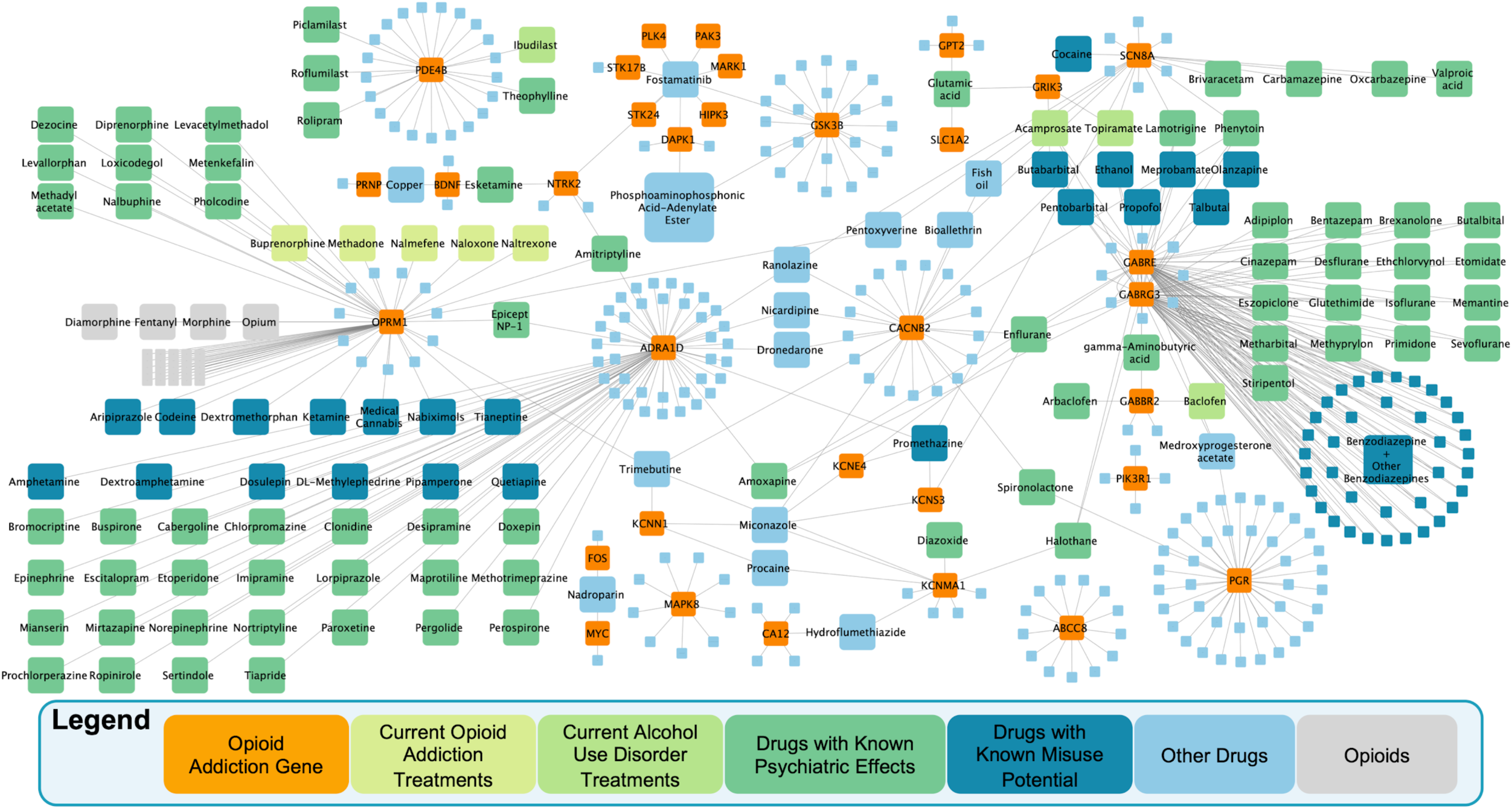
Drug-gene target network includes current opioid treatments and putative candidate drugs for treatment repurposing. A network of 414 drugs and 48 target genes (orange) implicated in multi-omic opioid addiction studies. Current opioid addiction/opioid use disorder (OUD) treatments (light yellow-green) targeting the μ-opioid receptor (*OPRM1*) include buprenorphine, methadone, and naltrexone, as well as nalmefene and naloxone to prevent overdose. In addition to nalmefene and naltrexone which is used to treat OUD and alcohol use disorder (AUD), four approved or experimental alcohol use disorder treatments (light green) were present in the network: acamprosate (targeting *GABRE* and *GABRG3*), baclofen (targeting *GABBR2*), ibudilast (targeting *PDE4B*), and topiramate (*CACNB2*, *GRIK3*, and *SCN8A*). Drugs with known psychiatric effects (teal) targeting opioid addiction genes include spironolactone (also investigated for AUD), antidepressants (*e.g.*, amitriptyline, amoxipine, and esketamine, targeting 6 unique genes). Some drugs in the network with known misuse potential (dark blue) include antipsychotic drugs (*e.g.*, aripriprazole, quetiapine) and many benzodiazepines that act as anxiolytics (*e.g.*, alprazolam, lorazepam). Multiple ion channel receptor subunits (15 total) are also known drug targets, and 9 genes are known to be targeted by fostamatinib. Other drugs with yet unknown psychiatric effects targeting opioid addiction genes are shown in light blue, along with a number of opioids (gray). This drug-gene target network may guide additional hypotheses and follow up experiments to test the efficacy of these drugs in combating opioid addiction processes.

In addition to drugs approved for treating substance use disorders, multiple drugs in the network targeting opioid addiction multi-omic genes are currently approved for psychiatric treatment or are known to have psychiatric effects. This includes the drug spironolactone (targeting *CACNB2* and *PGR*), which has recently been investigated for treating AUD^30^, and multiple approved antidepressants^31,32^ such as amitriptyline (*ADRA1D*, *NTRK2*, *OPRM1*), amoxapine (*ADRA1D*, *GABRE*, *GABRG3*), and esketamine (*BDNF, NTRK2*). The network also includes known drugs of abuse (*e.g.*, opioids, ethanol, promethazine) and widely prescribed drugs with potential for misuse, including antipsychotics (*e.g.,* aripriprazole^33,34^ (*OPRM1*), dosulepin^35^ (*ADRA1D*), olanzapine^33^ (*GABRG3*), pipamperone^33^ (*ADRA1D*), quetiapine^33,36^ (*ADRA1D*) and benzodiazepines (*e.g.*, alprazolam^37^, lorazepam^38^, temazepam^38^ that all target *GABRE* and *GABRG3*). A number of opioids are also present in the drug-gene target network based on their characterized effects on *OPRM1*.

Furthermore, multiple ionotropic and metabotropic ion channel and receptor subunits (*ADRA1D*, *CACNB2*, *GABBR2*, *GABRE*, *GABRG2*, *GRIK3*, *KCNE4*, *KCNMA1*, *KCNS3*, *NTRK2*, *OPRM1*, *SCN8A*) that were identified by multi-omic integration are known targets of approved drugs. Many drugs (“Other Drugs”, **Figure 5**) targeted 2 or more of the multi-omic opioid addiction genes that have not been investigated for psychiatric effects. For example, 9 genes (*DAPK1*, *GSK3B*, *HIPK3*, *MARK1*, *NTRK2*, *PAK3*, *PLK4*, *STK17B*, and *STK24*) were targeted by fostamatinib, indicating its potential to affect many opioid addiction genes simultaneously. Furthermore, we identified antagonists and agonists for most of the gene targets present in the conceptual model in **Figure 4** that were druggable, except for *FURIN* (only inhibitors), *GABBR2* (only agonists), *LGALS3* (unknown action), *SLC1A2* (substrate or inhibitors), and *SST* (only substrates). Thus, many of the genes in the biological pathways identified by our multi-omic integration are druggable targets and are novel candidates for drug repurposing studies.

## Discussion

In the present study, we integrated multi-omic datasets using a systems biology approach to identify biological pathways and drug targets underlying opioid addiction. Importantly, by integrating genome-wide association studies and omics derived from the dlPFC of individuals who died from an opioid overdose, we identified distinct biological pathways implicated in this brain region, which is associated with opioid craving^39–41^, such as the Akt and BDNF signaling pathways, which were not identified in the original contributing studies. Drug-gene target network integration identified candidate drugs targeting gene products implicated in opioid addiction omics, as well as putative gene targets for experimental follow up. These efforts may inform the development of therapeutic interventions for opioid addiction.

After identifying GWAS and omics-derived genes using consistent thresholds across opioid addiction studies, we used GRIN^17^ as a means of integrating these multi-omic data by identifying the most interrelated genes based on biological network connectivity. We used our previous validated multiplex network containing a dlPFC-derived predictive expression network in order to remove false positive genes from our gene set. Moreover, the dlPFC-specific network layers were congruent with the tissue of all transcriptomic and epigenetic data sets examined. Importantly, the 211 genes retained by GRIN were enriched for more GO Biological Processes than the 404 genes not filtered by GRIN. From the 211 genes, we identified a subset of 50 that were either previously implicated in the risk for substance use disorders, in opioid receptor signaling, or by multiple opioid addiction omics. Based on the high AUROC value from our random walk with restart cross-validation approach and the fact that only 127 genes were needed to connect opioid addiction genes in a shortest paths network, we concluded that these 50 genes were highly interconnected in our multiplex network and therefore part of the same biological pathways. Notably, while some of these genes have previously been implicated in rodent models of opioid addiction (*e.g.*, *FOS*^42–44^, *MYC*^45,46^) or opioid receptor signaling (*NTN1*^47^, *RTP4*^48,49^), many of the other genes are not well-characterized in relation to opioid addiction or from human postmortem brain tissue.

Gene set enrichment analysis revealed a strong enrichment for the BDNF, ERK, and Akt/PI3K pathways from our opioid addiction genes. Notably, the study describing the LC/MS proteomics data set and two RNA-seq data sets previously implicated the p38 and ERK MAPK pathways^11,12^. Here, we confirm this finding and expand the implication of the ERK MAPK pathway beyond the genes identified in prior studies. Animal models have shown increased ERK phosphorylation in the hippocampus^50^ and of the ERK scaffold protein PEA-15^51^. Decreased BDNF and associated epigenetic alterations have also been observed in the ventral tegmental area of postmortem tissue from human heroin addicts^52^. Moreover, the Akt/PI3K pathway has been implicated in differential gene expression from whole blood of subjects diagnosed with opioid use disorder^53^. However, to our knowledge this is the first study from the dlPFC in which multiple genes in the BDNF (*BDNF*, *NTRK2*) and Akt pathways (*GSK3B*, *PIK3R1*, *RASGRF1*) were implicated by opioid-induced transcriptomic and epigenetic alterations.

We also note that many of the 45 genes in our dlPFC opioid addiction conceptual model affect synaptic plasticity or neuronal signaling, and were largely associated with H3K27 hypoacetylation and/or decreased gene expression. Both excitatory receptor subunits (*CACNB2*, *GRIK3*) and inhibitory receptor subunits (*ABCC8*, *GABBR2*, *GABRE*, *GABRG3*, *KCNMA1*, *KCNN1*, *OPRM1*) were implicated in opioid addiction, and all except the *GABRE* subunit were associated with decreased expression or less active chromatin. Moreover, the EAAT2 glutamate transporter (*SLC1A2*) was implicated by H3K27 hypoacetylation, an astrocytic transporter responsible for removing glutamate from the synaptic cleft that has been implicated in bipolar disorder and schizophrenia^54^. Coupled with a net decrease in immediate early gene expression (*ARC, EGR1, EGR2, EGR4, ETS1, ETV5, FOS, MYC, NPAS4*), genes whose expression is induced by cellular activation^55–59^, our opioid addiction genes are associated with decreased neuronal activity in the dlPFC. Future studies are warranted to determine if decreased gene expression and H3K27 hypoacetylation occur equally within excitatory and inhibitory neurons in the dlPFC.

Extending GWAS findings for opioid addiction, here we integrated omics data in the dlPFC based on its role in impulsivity and drug craving in opioid addiction^39–41^. This brain region has also been implicated in cigarette^60^ and cocaine craving^61^. Moreover, randomized clinical trials have been conducted using transcranial magnetic stimulation over this brain region to reduce opioid craving^62,63^. Although omics from whole blood^53,64,65^ and other brain regions, such as the midbrain^66^, nucleus accumbens^13,67^, orbitofrontal cortex^68^, and striatum^69,70^ have been generated from individuals with a history of opioid addiction, focusing on GWAS and the dlPFC enabled us to examine genes from a brain region-specific perspective. Future studies should examine biological pathways involving NAc omics, particularly the extent to which these pathways overlap with dlPFC pathways. Furthermore, while the H3K27ac ChIP-seq data set integrated in this study were derived from NeuN-positive neurons^11^, omics derived from other cell types (*e.g.,* astrocytes, microglia, oligodendrocytes) and neuronal subtypes can inform cell type-specific epigenomic, transcriptomic, and proteomic changes caused by opioid addiction.

After identifying opioid addiction genes in the dlPFC, we sought to identify which genes were known targets of previously approved medications using network visualization. Notably, approved treatments for opioid addiction^71^ (buprenorphine, methadone, naltrexone) or overdose (nalmefene and naloxone) were present within our drug-gene target network. In addition to these opioid-related treatments, drugs for which there is evidence of efficacy in treating alcohol use disorder were present in the network, including acamprosate^27^, baclofen^27^, ibudilast^28^, and topiramate. Of these, topiramate has limited evidence in clinical trials for its capacity to treat opioid addiction^72^, and has been extensively assessed for treating alcohol use disorder^29^ and to a lesser extent cocaine^73^ addiction. In addition to medications for treating substance use disorders, we identified multiple drugs that are widely prescribed for treating psychiatric disorders that may be comorbid with opioid addiction, such as depressive and anxiety disorders and schizophrenia. Studies are warranted to evaluate the extent to which these drugs could be used to treat opioid addiction. As many of the identified genes were associated with decreased gene expression or hypoacetylated H3K72ac ChIP-seq peaks, ascertaining whether these drugs increase (agonist) or decrease (antagonist) the downstream activity of these protein-coding genes is critical. Furthermore, an important question for drug repurposing efforts is how opioid addiction affects the expression of the genes identified in the dlPFC.

In considering drugs to be repurposed for treating addiction, weighing the potential risks associated with them is essential. A number of drugs in the network that are widely used in psychiatry, such as the benzodiazepines alprazolam^37^ and lorazepam^38^, have the potential for misuse; thus, any efforts to repurpose these drugs must consider their potential for adverse effects. Future work might also include using real world data to determine safety and whether there is evidence that the use of these medications is associated with decreased use of opioids.

There are limitations to the current study that are worth noting. Notably, the omics from the dlPFC included in the study are from bulk tissue, rather than individual cellular populations. As they become available, integrating single-cell transcriptomic and epigenetic datasets would give valuable insight into the neuronal and/or glial cell populations affected by opioid addiction in the prefrontal cortex. Adding single cell-specific context into networks (*e.g.*, including a glutamatergic neuron-specific predictive expression network) would provide additional cellular perspective into the relationships among the multi-omic genes, beyond a tissue-level perspective. Furthermore, gene dysregulation in the dlPFC was identified by comparing opioid overdose death cases to controls. Such differences in gene regulation may be attributable to a variety of causes (e.g., chronic opioid exposure / addiction, genetic risks for opioid addiction, acute death from an opioid overdose, differences in diet, other drug use, or circadian disruption). It is likely that functional studies in model organisms will be needed to differentiate such causes. Finally, while we have included currently available genes implicated by GWAS and omics data, there are likely additional genes contributing to opioid addiction that have not yet been implicated due to these genes failing to reach statistical significance. Thus, as sample sizes continue to increase from opioid addiction GWAS and postmortem omics data from opioid overdose, there may be additional genes and pathways implicated within the dlPFC based on increased statistical power.

In summary, we used network biology techniques to integrate multiple opioid addiction omics data sets from a systems biology perspective. By identifying biological pathways dysregulated in the dlPFC following opioid addiction and druggable gene targets in these pathways, we identified candidate medications that merit experimental follow up as potential treatments for opioid addiction.

## Online Methods

### Integrating opioid addiction multi-omic genes

Opioid addiction genes were integrated from the following types of previously published omics data sets involving postmortem dorsolateral prefrontal cortex (dlPFC) from control subjects and subjects who died of an opioid overdose: H3K27ac chromatin immunoprecipitation sequencing (ChIP-seq^11^), DNA methylation^15^, liquid chromatography-mass spectrometry (LC/MS^12^), and bulk RNA sequencing (RNA-seq^11–14^). We also included genes identified from opioid addiction genome-wide association studies (GWAS). Together, 404 unique genes present in the multiplex network were identified based on the following thresholding procedures.

Differential ChIP-seq peaks were derived from previous H3K27ac ChIP-seq obtained from prefrontal cortex samples of controls and subjects that died of opioid overdose (**Supplementary Table 1**)^11^. From 388 differentially acetylated H3K27ac ChIP-seq peaks based on a threshold of Bonferroni-corrected *p* < 1e^-7^, peaks were re-mapped to genes using the “annotatePeak” function from the ChIPseeker R function^74^ using hg19/GRCh37 coordinates and a 1kb window around the transcriptional start site to define the promoter region. This annotation was used in contrast to the GREAT^75^ peak-to-gene annotation listed in the original publication^11^, as ChIP-seeker annotates the location of peaks (i.e. intronal, exonal, promoter, 5’UTR, 3’UTR, distal intergenic) and mapped peaks to genes that GREAT did not annotate. Reciprocally, if ChIPseeker did not map a peak to a gene, the GREAT peak-to-gene annotation was used. Together, these 388 H3K27ac ChIP-seq peaks mapped to 267 unique genes. Furthermore, we included the 5 genes (*ASTN2*, *DUSP4*, *ENOX1*, *GABBR2*, *KCNMA1*) associated with variant enhancer loci lost in opioid overdose subjects using promoter-capture Hi-C that were statistically significant at FDR-corrected *p* < 1e^-6^, which added 2 unique genes to the H3K27ac ChIP-seq-associated gene list. Thirteen non-coding RNAs identified by ChIP-seq peaks were excluded from downstream analysis as they were not present in any layer of the multiplex network (**Supplementary Table 7**): *MIR30D*, *MIR3201*, *MIR3914-1*, *MIR4262*, *MIR4264*, *MIR4319*, *MIR4432*, *MIR4480*, *MIR4689*, *MIR4714*, *MIR4790*, *MIR488*, and *SNORA72*. In total, 256 unique genes were included among the H3K27ac ChIP-seq peaks.

Differentially methylated genes were identified using a previously published epigenome-wide association study of dlPFC tissue from subjects who died from opioid overdose using the Illumina Infinium MethylationEPIC BeadChip DNA methylation chip testing 850,000 CpG regions (**Supplementary Table 2**)^15^. Thirteen CpG sites were identified with adjusted *p*-values of 0.4 (uncorrected *p* < 6e^-6^) and 3 CpG islands that were not mapped to genes (cg25084741, cg10759972, and cg26506680) were excluded. This yielded 13 unique genes identified by opioid-induced differential DNA methylation that were present in the multiplex network.

Genes were included from opioid use disorder^8–10^ and prescription opioid misuse^6^ GWAS based on SNP-nearest gene associations from SNPs with *p* < 5e^-8^, resulting in 19 unique GWAS genes; an additional 15 unique genes were also identified from multi-trait analysis of GWAS (MTAG) using European opioid use disorder GWAS, alcohol use disorder, and cannabis use disorder summary statistics (**Supplementary Table 3**)^7^. These opioid addiction GWAS yielded 34 unique genes, but one gene (TMX2-CTNND1) was not present in any layer of the multiplex and was excluded from downstream analyses (33 unique GWAS genes; **Supplementary Tables 6-7**).

Differentially abundant proteins (**Supplementary Table 4**) were identified from a previously published LC/MS proteomics study from postmortem dlPFC from control and subjects diagnosed with opioid use disorder^12^. Three protein coding genes (*ATP5J2*, *LGALS3*, *TAF15*) were included based on a threshold of adjusted *p*-value < 0.05 and absolute value of log_2_ fold change (FC) greater than or equal to 1.5 (log_2_FC ≤ -1.5 or log_2_FC ≥ 1.5) which were included in downstream analyses.

Differentially expressed genes (DEGs; **Supplementary Table 5**) were identified from four RNA-seq studies derived from postmortem dlPFC of controls and subjects who died of opioid overdose or were diagnosed with opioid use disorder^11–14^. DEGs were included in downstream analyses based on thresholds of adjusted *p*-value < 0.05 and |log_2_FC ≥ 0.5|. Using these thresholds, we identified 106 unique genes identified in opioid addiction by RNA-seq and 103 genes present in the multiplex were included in downstream analyses (*AC018647.3*, *AL606753.2*, and *PARTICL* were excluded, **Supplementary Table 7**).

Multiple genes were implicated in opioid addiction by multiple lines of biological evidence. Four genes were identified across multiple omics data types: *DUSP4*, *DUSP6*, *ETV5*, and *PLA2GS*. Additionally, 60 ChIP-seq opioid addiction genes were identified by multiple H3K27ac ChIP-seq peaks, and *ARL4D* was identified as a DEG in two separate RNA-seq studies^12,13^. Furthermore, *FURIN* ^7–9^, *NCAM1* ^7,8^, *OPRM1* ^7–10^, and the *SCAI*/*PPP6C*/*RABEPK* ^7–9^ gene cluster were identified across multiple opioid addiction GWAS.

### Multiplex network generation

Multiplex gene-gene networks were assembled from 10 layers from different types of biological evidence using GENCODE IDs for all genes. None of the multiplex network data sources were from any of the opioid addiction omics data sets. The following network layers were used from HumanNet (version 2.0 ^23^): co-citation, co-essentiality, co-expression, pathway databases, gene neighborhood, interologs, and phylogenetic associations. A merged protein-protein interaction network was created by merging networks from HumanNet (version 2.0, literature curated and high-throughput assay-derived connections) and high-confidence interactions from STRING (version 11.0, taxa = 9606, protein.actions.v11.0, mode=binding, min score = 700^76^). Transcription factor-target relationships specific to the prefrontal cortex were included from Pearl et al^20^. All edge (connection) weights were normalized on a 0 to 1 scale irrespective of line of biological evidence.

Brain region-specific network layers were also incorporated using RNA-seq data from the Genotype-Tissue Expression (GTEx) consortium^77^ and explainable-AI algorithms applied on the Summit supercomputer at the Oak Ridge Leadership Computing Facility (OLCF). Using iterative random forest leave-one-out prediction (iRF-LOOP^78^), gene-gene predictive networks were made from the prefrontal cortex (BA9) given that all omics were derived from this brain region. After computing all edges with iRF-LOOP, only high-confidence edges (edge weights > 0.05) were included in the multiplex network.

After compiling all network layers, the 10-layer multiplex was assembled using the “RWR_make_multiplex” function within the RWRtoolkit R library^79^. The final multiplex network contained 51,183 unique genes and 3,419,975 unique edges.

### Filtering opioid addiction multi-omic genes using GRIN

We applied GRIN (Geneset Refinement using Interacting Networks)^17^ to identify the most biologically interrelated opioid addiction genes and remove potential false positive genes. GRIN was applied with the previously validated 10-layer multiplex network described above using the 404 unique genes identified from multiple omics data using equal weights for all genes. GRIN retained 211 opioid addiction genes which were used in all subsequent analyses. Gene set enrichment for Gene Ontology (GO) Biological Processes was then tested on the 404 genes prior to GRIN as well as the GRIN retained and GRIN removed gene sets using ToppGene^80^. Gene set enrichments were considered significant at a threshold of FDR-corrected *p*-value < 0.05.

After retaining 211 opioid addiction genes, a set of 50 high-confidence genes were identified using two criteria. Three genes that were present in multiple omics data sets and retained by GRIN were included: *DUSP4*, *DUSP6*, and *ETV5*. Next, a subset of the 211 genes was included if each gene were associated with opioid-mediated signaling or a substance use disorder based on previously published research. This resulted in 47 additional genes in the high-confidence gene set based on a literature search from PubMed (https://pubmed.gov) using the following search terms: “[gene] opioid”, “[gene] addiction”, “[gene] substance use disorder.” ToppGene was then used to test for gene set enrichment of GO Biological Processes, GO Cellular Components, GO Molecular Functions, biological pathway enrichment (includes KEGG^81^, Reactome^82^, and PANTHER^83^ pathways), and transcription factor binding sites (MSigDB^84^).

### Shortest paths network

A shortest paths network connecting all pairs of the 50 opioid addiction-associated genes was generated using the shortest paths algorithm using the R package RWRtoolkit^79^. The 10-layer multiplex network was merged into a single network layer, and using the shortest paths algorithm identified the shortest possible network connections among all 2500 possible gene-gene pairs. Genes who were direct neighbors with at least one other opioid addiction gene (43 out of 50 opioid addiction genes) were imported into the program Cytoscape^85^ (Version 3.9.1, Cytoscape Consortium, San Diego, CA, USA) for network visualization. For the other 7 genes (*ADRA1D*, *FURIN*, *KIAA0040*, *LGALS3*, *NTN1*, *SLC1A2*, *SST*), the connections between these genes and other opioid addiction genes that were separated by one neighbor (“network-connecting genes”) were included in the Cytoscape visualization.

### Random walk with restart cross validation

Using the RWR-CV function within RWRtoolkit^79^, we tested the interconnectivity of our refined 50 opioid addiction gene set using random walk with restart (RWR) with 5-fold cross validation. In one cross validation fold, 10 genes were left out of our input gene set, and the other 40 genes were used as seed genes for RWR. All genes in the multiplex network were ranked by RWR, with true positives counted as left-out genes that were ranked by RWR, and true negatives counted as other genes in the network. A receiver-operator characteristic (ROC) curve was calculated based on true positive rate (true positives / true positives + false negatives) and false positive rate (false positives / false positives + true negatives), which was plotted using the R package ggplot2^86^. The area under the curve was computed using RWRtoolkit.

### Overview and conceptual model visualizations

An overview diagram of the approach used in this study was constructed using Biorender.com. Similarly, the conceptual model illustrating biological pathways among opioid addiction genes was constructed using Biorender.com.

### Identifying druggable targets involved in opioid addiction

Multi-omic opioid addiction genes were then cross-referenced with previously demonstrated gene-drug interactions. Using DrugBank^87^, a network of drug-gene target interactions was created using all 211 genes retained by GRIN. These drug interactions included FDA-approved, investigational, and experimental uses of drugs targeting these genes, resulting in 48 gene targets and 414 total drugs. Drug-gene target interactions were visualized using Cytoscape.

## Data Availability

No primary data was generated for the present study. All primary data from DNA methylation, GWAS summary statistics, H3K27ac ChIP-seq, LC/MS proteomics, and RNA-seq are from previously published manuscripts. GWAS summary statistics from the Million Veteran Program (Kember et al., 2022) are available on the NIH database of Genotypes and Phenotypes (dbGaP) under accession phs001672. GWAS summary statistics from Deak et al., 2022 are publicly available at https://medicine.yale.edu/lab/gelernter/stats/. GWAS summary statistics from Gaddis et al., 2022 are available under dbGaP under accession phs000454.v1.p1.We used the top 10,000 SNPs from Sanchez-Roige et al., 2021 GWAS summary statistics, which are publicly available at https://www.ncbi.nlm.nih.gov/pmc/articles/PMC8562028/bin/41380_2021_1335_MOESM2_ES M.xlsx. Previously published data from H3K27ac ChIP-seq is available under dbGaP accession number phs002724.v1.p1. Previously published DNA methylation data is available at GEO accession number GSE164822. Previously published LC/MS proteomics is available at the ProteomeXchange PRIDE repository under PXD025269. Previously published RNA-seq data sets are available under dbGaP accession phs002724.v1.p1, GEO accession numbers GSE221515 and GSE174409, and SRA accession number SUB9455518.

All activities were approved by the Oak Ridge National Laboratory Institutional Review Board. The demographics of subjects from which GWAS summary statistics were derived along with descriptions of Institutional Review Boards to approve these studies have been characterized in previous publications and all subjects provided informed consent. All postmortem brain tissue samples are exempt from human subjects research.

## Code Availability

The GRIN software and multiplex network that was used is publicly available at github.com/sullivanka/GRIN. Publicly available R packages (ggplot2, tidyverse) were used for data analysis and visualization using R version 4.1.3, and ChIP-seq peaks were assigned using ChIPseeker (version 1.30.3). Additional code used to generate results are available upon request.

## Acknowledgements/Funding Sources

This manuscript has been authored by UT-Battelle, LLC under Contract No. DE-AC05-00OR22725 with the U.S. Department of Energy. The United States Government retains and the publisher, by accepting the article for publication, acknowledges that the United States Government retains a non-exclusive, paid-up, irrevocable, world-wide license to publish or reproduce the published form of this manuscript, or allow others to do so, for United States Government purposes. The Department of Energy will provide public access to these results of federally sponsored research in accordance with the DOE Public Access Plan (http://energy.gov/downloads/doe-public-access-plan). This research used resources of the Oak Ridge Leadership Computing Facility, which is a DOE Office of Science User Facility supported under Contract DE-AC05-00OR22725. This work was funded by NIH grants DA051908 (EOJ, DAJ), DA051913 (DBH, DAJ), and DA054071 (NCG, OC, BSM, SS, AP, VT, EC, EOJ, DAJ) and VA grant I01 BX004820 (ACJ, HRK) and the Veterans Integrated Service Network Mental Illness Research, Education and Clinical Center (HRK). This research is based on data from the Million Veteran Program, Office of Research and Development, Veterans Health Administration, and was supported by award # I01 BX004820. This publication does not represent the views of the Department of Veteran Affairs or the United States Government.

## Author Contributions

Conceptualization: DBH, EOJ, DAJ; Methodology: KAS; Software: KAS, DK, ML, MC, JIM; Formal analysis: KAS; Investigation: EOJ; Writing - Original Draft: KAS, EOJ; Writing - Review & Editing: KAS, MRG, AT, BCQ, CW, NCG, RM, OC, BSM, PCS, SS, AP, VT, EJC, RLK, HRK, ACJ, KX, BEA, DBH, EOJ, DAJ; Supervision: DBH, EOJ, DAJ; Funding acquisition: HRK, ACJ, DBH, EOJ, DAJ.

